# Evaluation of Machine Learning and Traditional Statistical Models to Assess the Value of Stroke Genetic Liability for Prediction of Risk of Stroke within the UK Biobank

**DOI:** 10.1101/2025.02.21.25322620

**Authors:** Gideon MacCarthy, Raha Pazoki

## Abstract

**Background and objective:** Stroke is one of the leading causes of mortality and long-term disability in adults over 18 years of age globally and its increasing incidence has become a global public health concern.

Accurate stroke prediction is highly valuable for early intervention and treatment. Previous studies have utilized statistical and machine learning techniques to develop stroke prediction models. Only a few have included genome-wide stroke genetic liability and evaluated its predictive values. This study aimed to assess the added predictive value of genetic liability in the prediction of the risk of stroke.

**Materials and methods:** The study included 243,339 participants of European ancestry. Stroke genetic liability was constructed using previously identified genetic variants associated with stroke by the MEGASTROKE project through genome-wide association studies (GWAS). In our study, we built four predictive models with and without stroke genetic liability in the training set: Cox proportional hazard (Coxph), Gradient boosting model (GBM), Decision tree (DT), and Random Forest (RF) to estimate time-to-event risk for stroke. We then assessed their performances in the testing set.

**Results:** Each unit (standard deviation) increase in genetic liability increases the risk of incident stroke by 7% (HR = 1.07, 95% CI = 1.02, 1.12, P-value = 0.0030). The risk of stroke was greater in the higher genetic liability group, demonstrated by a 14 % increased risk (HR = 1.14, 95% CI = 1.02, 1.27, P-value = 0.02) compared with the low genetic liability group. The Coxph model including genetic liability was the best-performing model for stroke prediction achieving an AUC of 69.54 (95% CI = 67.40, 71.68), NRI of 0.202 (95% CI = 0.12, 0.28; P-value = 0.000) and IDI of 1.0×10^-04^ (95% CI = 0.000, 3.0×10^-04^; P-value = 0.13) compared with the Cox model without genetic liability.

**Conclusion:** Incorporating genetic factors in the model may provide a slight incremental value for stroke prediction beyond conventional risk factors.

## 1. Introduction

Stroke is one of the leading causes of mortality and long-term disability in adults over 18 years of age globally and its increasing incidence has become a global public health concern [1, 2]. In addition, stroke survivors have a considerably higher risk of mortality when compared with non-stroke patients, not only attributed to the initial stroke but also to stroke-associated consequences and increased cardiac incidence in years after a stroke [3–6]. Globally, the rising incidence of stroke is and will be a cause of concern due to its detrimental impact on the economy and the increasing cost of healthcare and social services throughout the world. Every year, more than 100,000 people in the United Kingdom (UK) suffer from a stroke, and over 1.2 million stroke survivors live in the UK. Stroke incidence and prevalence in the UK are expected to increase by 60% and 120% annually between 2015 and 2035, respectively [7].

Both genetic and non-genetic risk factors contribute to the complex mechanism of stroke [8]. The risk of stroke increases with age, and the estimated 10-year stroke risk in adults aged 55 and over. The risk differs by sex and the increasing co-occurrence of risk factors such as hypertension, DM, atrial fibrillation, high blood cholesterol and lipids, cigarette smoking, physical inactivity, chronic kidney disease, and family history [9].

Twin and family history studies provided early evidence that genetics had a role in stroke risk [10]. Genome-wide association studies (GWAS) have provided further evidence to confirm the role of genetic factors in the occurrence of stroke.

More recently, large-scale GWAS such as the International Stroke Genetics Consortium (ISGC) have identified genetic loci associated with stroke. The MEGASTROKE project identified over 32 loci contributing to stroke risk, revealing the causal role of specific genes and gene regions in stroke origins [11, 12]. As a result, greater insight into the genetic indicators of stroke has allowed an opportunity for a deeper evaluation of an individual’s stroke risk, as well as potentially more informed medical and lifestyle decisions that may be preventative measures to reduce the risk of stroke occurrence.

Prediction tools for stroke, such as the Framingham Stroke Risk Profile (FSRP) the American Heart Association (AHA), and the American Stroke Association (ASA), are critical in identifying at-risk individuals early on, allowing for timely treatments and improving outcomes [13, 14]. Their developments extend beyond individual treatment, including healthcare policy, budget allocation, and ethical issues for patient data. Advances in artificial intelligence and machine learning are pushing the boundaries of prediction tools, making them more accurate and adaptive to diverse groups of patients [15].

The stroke prediction tools (FRSPs and ASA), as well as the current clinical guidelines for cardiovascular disease prevention, do not evaluate or integrate genetic liability into the risk assessment [16].

Genetic prediction of stroke has the potential to transform stroke prevention and treatment. It has the potential to identify individuals who are at risk of or predisposed to stroke even before clinical symptoms appear. This allows for early treatments such as lifestyle adjustments or personalized drug programs [17–21].

Genetic polymorphisms in genes associated with stroke or its risk factors have been investigated in stroke risk, with several studies reporting that genetic liability derived from a set of SNPs with a strong association with stroke or stroke risk factors had a significant association with stroke risk [22–28].

Genome-wide genetic liabilities, derived from the combined effects of several genetic variants across the genome, regardless of the strength of their association, have been increasingly tested in the last decades for their effect in health and disease, and several studies have shown that higher scores of genome-wide genetic liabilities enhance the stroke risk prediction [29, 30].

Machine learning models are increasingly applied to predict the risk of complex diseases [31–42]. Studies focusing on the prediction of the risk of stroke [32, 41, 42] have shown that machine learning models outperformed traditional statistical techniques such as the Cox proportional hazards model. However, there is no consistency on which machine learning model is a better fit. Chen et al. [42] identified Artificial Neural Network (ANN), whilst Chun et al. [32] found Gradient-Boosted trees (GBT) and Wang et al. [41] identified Random Forest outperforming the Cox proportional hazards model.

In addition, the predictive value of genetic factors used in machine learning models is unclear. In a case-control study focusing on patients with atrial fibrillation, Papadopoulou et al. [40] showed that out of multiple machine learning models incorporating a genetic liability, XGBoost outperformed a widely used existing clinical prediction model (CHA2DS2-VASc). The study by Papadopoulou et al. [40] did not include incident stroke and they created their genetic liability using a selected list of single nucleotide polymorphisms (SNPs) associated with ischemic stroke.

To our knowledge, there is currently no study in the European general population to provide a comprehensive insight into the prediction of risk of incident stroke in various scenarios incorporating machine learning and a stroke genome-wide genetic liability. To fill this gap, our research focussed on incorporating a genome-wide genetic liability into machine learning for the prediction of the risk of incident stroke using survival data. This would offer a better understanding of the additional benefit of genetic liability in stroke risk prediction, as well as how machine learning algorithms perform in comparison to traditional survival models in this context.

We have three main objectives including (1) assessing the association of whole genome liability and the risk of future stroke occurrence (incident stroke), (2) assessing the predictive value of stroke genetic liability in the prediction of stroke (3) comparing the performance of the Cox proportional hazard model and machine learning models before and after incorporating genome-wide stroke genetic liability into the model.

## 2. Material and Method

### 2.1. Ethical Approval

The Northwest Multi-Centre Research Ethics Committee gave ethical approval to the UK Biobank (UKB) as a research tissue bank, and each participant gave informed consent. The current study is conducted on UKB data under application number 60549. In addition, we received ethical permission from Brunel University London’s College of Medicine and the Life Sciences Research Ethical Committee to work with UKB secondary data (reference 27684-LR-Jan/2021-29901-1).

### 2.2. Study Population

UKB is a prospective observational study with over 500,000 participants aged 40 to 69. Between 2006 and 2010, participants were recruited from 22 centers across the United Kingdom. The comprehension description of the UKB study, the data obtained, and a summary of the characteristics are publicly available and can be accessed on the UKB website (www.biobank.ac.uk, accessed on 20 June 2021) and elsewhere by Sudlow and colleagues [43]. Detailed information about socio-demographics, health status, physician-diagnosed diseases, family history, and lifestyle variables was obtained during the recruitment phase using questionnaires and interviews. A variety of physical measurements were taken, including height, weight, body mass index (BMI), waist-hip ratio (WHR), systolic blood pressure (SBP), and diastolic blood pressure (DBP). The UKB experiment participants’ records were linked to the health episode statistics (HES) data, as well as national death and cancer registries.

The current study is focused on a subset of unrelated individuals of European ancestry (N = 243,399; **Figure 1**). In summary, we used 40 genetic principal components created centrally by the UKB and the k-means clustering approach on 502,219 UKB participants to identify people of European ancestry (N = 459,042) and have genetic data. Participants who had withdrawn their consent (N = 61), pregnant women, and those unsure of their pregnancy status (N = 278) were excluded from the study. We also excluded participants with discordant genetics and self-reported sex (n = 320). Using a kinship cut of 0.0884 for third-degree relatives, we excluded individuals who were up to second-degree related (N = 33,369). We excluded participants (N = 25,340) who had been diagnosed with vascular/heart problems by a doctor before or during recruitment. Participants who were using cholesterol-lowering medication (N = 34,243), ceased smoking, or drinking due to health reasons or doctor’s recommendation (n = 58,752), and had missing data on confounders (N= 61,961) were also excluded from the dataset. We subsequently excluded participants who had prevalent stroke cases (N= 248), and self-reported stroke (N=130). We subsequently merged the data with genetic liability profile data (N = 425,054) calculated for participants with available genotype data (N = 459,042), leaving a final 243,399 unrelated individuals of European ancestry.

**Figure 1.**
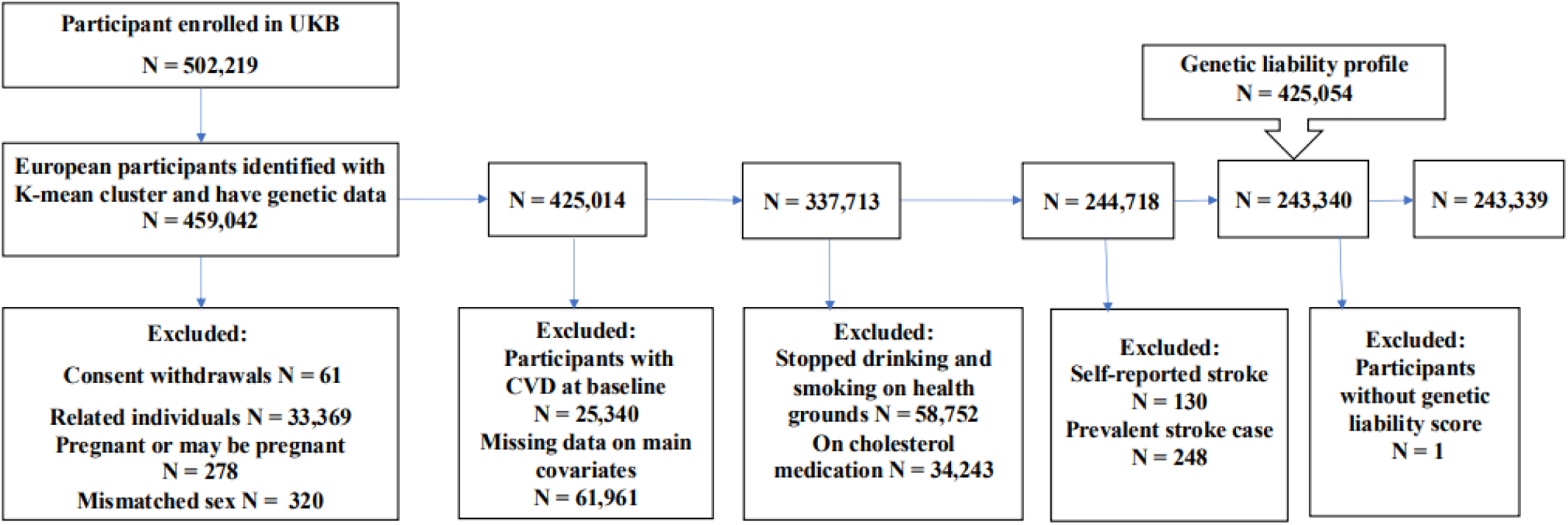
Exclusion Criteria of the study: The flowchart of the study participant selection. UK Biobank (UKB) data had over 500,000 participants at the time of the beginning of this study. We used the K-mean cluster method to extract 459,042 participants of European ancestry and have genetic data. The final dataset included 243,339 participants who met the inclusion criteria.

### 2.3. Genotyping and Imputation

All DNA extraction, genotyping, and imputation were conducted by the UKB. Detail techniques have been described elsewhere [44–46]. In summary, blood samples from participants were collected at UKB assessment centers, and DNA was extracted and genotyped using the UKB Axiom array. The genotyping imputation was performed by UKB using the IMPUTE4 program [47]. The imputation was done using three reference panels: the Haplotype Reference Consortium, UK10K, and 1000 Genomes Phase 3. The UKB centrally estimated the genetic principal components and kinship coefficients to account for population stratification and identify related individuals [44, 46].

### 2.4. Definition of the Outcome

Our primary outcome in the current study was stroke events, defined according to the International Classification of Diseases 10th revision (ICD-10, I60– I67). In this study, incident stroke was characterized using cerebrovascular disorders ICD-10 code (I600-I609, I610-I619, I630-I639, I64, I650-I659, I660-I669, and I670-I679) for the first stroke event. The current study’s follow-up period is computed from the date of health assessment upon enrolment to the end of March 2017. The participants who did not experience the outcome at the end of the follow-up period were censored.

### 2.5. Demographics, Clinical and Lifestyle Features

In all analyses, conventional risk factors such as age, sex, BMI, diabetes mellitus (DM), hypertension, total cholesterol (TC), low-density lipoprotein (LDL), smoking status, and drinking status, were considered as demographic, clinical and lifestyle features. We defined DM as a record of diabetes diagnosed by a doctor, the use of insulin treatment, or the blood level of haemoglobin (HbA1c) ≥ 48 mmol/mol (6.5%), or glucose level ≥ 7.0 mmol/dL [48]. We define hypertension as (1) the presence of a recorded SBP ≥ 140 mmHg or a DBP ≥ 90 or (2) hypertension diagnosed by a doctor or (3) a record of using blood pressure (BP) lowering medication at baseline [49, 50].

In the UKB, two blood pressure measurements were taken a few minutes apart using a standard automated instrument or a manual sphygmomanometer (https://biobank.ctsu.ox.ac.uk/ukb/ukb/docs/Bloodpressure.pdf). We estimated mean SBP and mean DBP based on two automated or two manual blood pressure measurements. For individuals with one manual and one automatic blood pressure reading, the average of the two results was used. For individuals having a single blood pressure measurement (either manual or computerized) that single measurement was used. We increased SBP by 15 mmHg and DBP by 10 mmHg for individuals who self-reported taking blood pressure-lowering medications [51]. Participants with missing blood pressure values were excluded. Smoking and alcohol consumption data were obtained through a self-reported questionnaire by the UKB and were classified into current, previous, and never.

### 2.6. Computation of Genetic Liabilities

#### Single nucleotide polymorphisms (SNP) Selection

We selected a list of genetic variants in the form of SNPs (Supplementary Data S1) that were previously discovered as associated with stroke [52]in the European population. The effect sizes (Supplementary Data S1) were obtained from previously published, publicly accessible GWAS summary statistics data for these SNPs on the GWAS Catalog website (https://www.ebi.ac.uk/gwas/, viewed on July 12, 2021).

For this study, we excluded duplicate, non-biallelic SNPs and SNPs with a minor allele frequency (MAF) less than or equal to 0.01 from the process of calculating the genetic liability. We further performed a linkage disequilibrium (LD) pruning procedure to exclude SNPs that are in LD with each other.

SNPs are in LD when the association between their alleles occurs more often than expected in a random sample [53]. LD between two loci is calculated statistically using parameters such as r^2^ value. This metric measures the level of correlation between alleles at the two loci. In brief, LD pruning ensures that redundancy is reduced by removing highly correlated SNPs. This is done to avoid statistical bias and computational inefficiency caused by LD.

The LD pruning procedure based on correlation coefficient (r^2^) removes SNPs with high LD by calculating r2 between pairs of SNPs within a given frame. For this LD pruning process, all pairs of SNPS within a given moving window are evaluated to determine their pairwise LD. If any pair of SNPs within the window has LD larger than the stated threshold, the first SNP will be pruned [54].

In this work, we used r^2^ = 0.1 as the threshold for pruning SNPs that are in LD with others. The pruning process was implemented in PLINK version 1.9 [55] with the function and parameters *“- -indep-pairwise window size = 250 step size = 50 r^2^ = 0.1”.* After the LD pruning procedure, 252,903 SNPs were retained for calculating the genetic liability for stroke (**Supplementary Figure 1**). The calculation of genetic liability for stroke was implemented in PLINK version 1.9 with the function “*-score”.* PLINK employs a weighted technique in which the effect size (beta coefficient) of each SNP is used as weight and is multiplied by the number of risk alleles carried by the participant. The result is then summed up across all SNPs in the calculation of genetic liability.

### 2.7. Data Preprocessing

We pre-processed the dataset by standardizing all quantitative variables including age, BMI, LDL, and genetic liability using the “*scale”* function in the R package. Categorical variables included sex (male and female), smoking status (never, previous, current), alcohol consumption status (never, previous, current), DM (no, yes), and hypertension (no, yes). Genetic liability was additionally categorized as low, medium, and high risk according to its tertiles to ease analysis per subgroup of genetic liability.

### 3.0. Statistical Analysis

For a statistical description of the baseline characteristics of our study population, we used the *“gtsummary”* and “*table1*” packages in the R-program Windows version 4.4.1 for statistical analyses [56]. The categorical variables were summarized using frequencies and percentages and the numerical variables were expressed as mean (SD). The Chi-square test was used to compare differences in binary outcome (stroke event and non-event) in relation to categorical variables. For continuous variables, the Wilcoxon rank sum test was used. We used univariable Cox proportional hazard regression to assess the association between each non-genetic feature and the outcome. We also assessed the collinearity between the numerical features to identify features that are highly correlated with one another. We used the *“cor”* function to calculate the correlation matrix and the Pearson correlation between variables and the *“ggcorrplot”* function from the *ggcorrplot* package to visualize the correlation matrix. We then examined the correlation matrix using the *“findCorrelation*” function from the *caret* package to identify highly correlated features. In this study, we set the Pearson correlation (r^2^ = 0.8) as the threshold for collinearity [57, 58]. We excluded the features highly correlated with one another (r^2^ greater than 0.8) and those with a weaker association with stroke as indicated by a smaller alpha error.

### 3.1. The Relationship Between Genetic Liability and Stroke

We used univariable and multivariable Cox proportional hazard regression to assess the relationship between stroke genetic liability (continuous and categorical) and the risk of incident stroke over the follow-up period.

Hazard ratios (HR) are commonly used to evaluate outcomes like survival time, and time to event. HR is a measure used in survival analysis to compare the risk of an event occurring at any given point in time between two groups.

Following the univariable Cox proportional hazard regression analysis (model 1), three multivariable adjustment Cox proportional hazard regression models (models 2, 3, and 4) were developed to examine the potential influence of known cardiovascular risk factors on the relationship between genetic liability and stroke risk. In model 2, we adjusted for age and sex. In model 3, BMI, hypertension, DM, and LDL were adjusted in addition to age and sex, and in model 4, we further adjusted for drinking status and smoking status (Full model). We defined statistical significance when the associations establish a 2-sided P-value less than 0.05. We assessed the proportional hazard (PH) assumptions using statistical testing (*cox.zph”* function) and visual examination of scaled Schoenfeld residuals (*“ggcoxzph”* function) using the R *survival* package.

### 3.1 Prediction Models Development

In this study, two sets of prediction models were created for each technique to predict the incidence of stroke. (1) The conventional risk factors model (model without genetic liability), which combines the conventional risk factors selected from univariable association tests and (2) The integrated prediction model, which combines genetic liability for stroke (genetic risk) and the conventional risk factors.

Using the “*createDataPartition*” function from the *caret* package, we randomly partitioned our dataset into a training set (70%; N = 170,381; Event = 1,382 and Non-event = 168,999) and a testing set (30%; N = 73,018; Event = 591 and Non-event = 72,427).

To predict the risk of incident stroke, we used the training data to create prediction models using the Cox proportional hazard. Cox proportional hazards regression [59, 60] is a popular statistical approach for assessing survival data and determining the association between the time until an event (such as death, failure, or illness recurrence) occurs and one or more predictors. We implemented the Cox proportional hazard models using the *“coxph”* function from the *Survival* package in R software.

In addition, we developed three machine learning techniques in the training set including the Gradient Boosting Machine models (GBM), Decision Tree (DT), and Random Forest (RF) to predict the risk of stroke. We then assessed the performance of each model in the testing set (Supplementary Figure 2).

The Decision Tree is one of the common and simple methods used for classification and regression applications. It works by dividing a dataset into smaller subgroups depending on feature values and then generating a decision tree [61]. The Decision Tree method in this study was implemented using the “*rpart”* function from the Recursive Partitioning and Regression Trees *(rpart*) package, the minimum number of observations required to split a node at each branch was set to 4. The complexity parameter (cp) to control the size of the decision tree and prevent overfitting was set at 0.001 meaning that a split must improve the model’s fit by at least 0.1% to be considered. This parameter is used to save computing time by removing irrelevant splits. The optimal decision tree was obtained with the *“prune”* function. The function removes the trees that do not meet the complexity parameter value. That is the *“prune”* function removes branches without lack of fit reduction (measured by the residual sum of squares; RSS) as determined by the complexity parameter value. This process reduces the risk of overfitting the training data.

Random Forest is a popular machine learning model for classification and regression. It creates ensembles from decision trees and combines their results to make a final decision [62]. The Random Forest models were built using the “*ranger”* function from the *ranger* package. The number of trees to be fitted was set to a value of 500. To control the model’s complexity and performance, the number of variables randomly selected at each split when growing the trees was set to a value of 3 (“*mtry*”). This is justified as the optimal *mtry* value considered for classification models is calculated as the square root of the total number of variables (9 variables in the current study). The value of *mtry* can significantly affect the OOB (out-of-bag) error. The OOB error is an unbiased estimate of the prediction error calculated by using samples not included in the bootstrap sample for a given tree. It serves as a cross-validation mechanism that is integrated into Random Forest. A smaller *mtry* value increases the randomness and diversity among the trees, which can help reduce overfitting and potentially lower the OOB error.

However, if the *mtry* value is too small, the trees might not capture enough information, leading to higher OOB error. The *mtry* and OOB error are critical in optimizing the random forest model. The range of values for *mtry* was examined by the *ranger* package and the *mtry* value that minimizes OBB error was selected as the optimal value in the construction of the Random Forest model. We additionally built Gradient Boosting Machine models using the *gbm* package to predict the risk of stroke. The Gradient Boosting Machine models integrate predictions from many weak learners to increase total prediction accuracy [61]. The number of trees to be fitted was set to a value of 500. The highest number of permissible variable interactions was set to 3. The shrinkage parameter to control the learning rate or step-size reduction was set to a value of 0.01. Lastly, the number of cross-validations (cv) performed was set to a value of 10.

### 4.0. Model Performance Assessment

To determine the confidence of prediction and the predictive performance of each model, we used the Platt-scaling method [63] also known as the sigmoid method, to calibrate the probability estimates provided as the output of the models. Platt scaling is a method used in machine learning techniques to transform the output from classification models into a probability distribution over classes. This was done by passing the probability estimates from machine learning models through a trained sigmoid function [63] using a univariable logistic regression in which the probability estimates were used as independent variables and the binary outcome (stroke) served as the dependent variable [64]. This provides new scaled probability estimates to help calibrate the models. Calibration of a prediction model ensures that the predicted risks are accurate and align with the actual proportions of the event. A prediction model is said to be calibrated if the model’s outcome matches the observed proportions of the event [65]. To assess model calibration, we used the *“pmcalibration”* function from *pmcalibration* in the R package within which restricted cubic lines, a type of smoothing function, was used (to create calibration curves for the prediction models). This method allows for nonlinear relationships between the predictors and the response variables. Complementary log-log transformed predicted probabilities were applied to the splines to produce calibration measures for a time-to-event outcome.

The calibration metrics used to assess the model calibration in this study were the Brier Score (*BS*) and average absolute difference (*Eavg*) also known as the integrated calibration index (*ICI*). BS is the mean squared difference between the predicted probabilities and the actual outcomes, and it measures both discrimination and calibration [64]. BS ranges from 0 (perfect prediction and calibration) to 1(worse prediction and calibration). ICI measures the average absolute deviation between the predicted and observed probabilities, providing an overall assessment of calibration quality [65]. It provides a single, summary measure of calibration quality, making it easier to compare different models or assess changes in calibration over time. An ICI of 0 represents perfect calibration and an ICI of 1 represents worse calibration, suggesting that the predicted probability deviates from the observed events. To calculate ICI and BS, we used “*pmcalibration*” and *“brier”* functions implemented within the *pmcalibration* and *gmish* packages, respectively.

To assess the discrimination performance of the models, we calculated the area under the curve (AUC) using the *pROC* package in the R program. We reported AUC, ICI and BS of various models. Greater values of AUC and smaller values of ICI and BS indicate improved discrimination and calibration of the model.

### 4.1 Assessment of the predictive value of genetic liability

We assessed the predictive value of genetic liability as an additional predictor to the conventional risk factors in each prediction model by estimating improvement in the AUC, Integrated Discrimination Improvement (IDI), and continuous Net Reclassification Index (NRI). NRI measures the effectiveness of a new model in reclassifying individuals into different risk categories compared to an existing model. At the same time, IDI evaluates the model’s ability to differentiate between cases and non-cases after adding a new variable. It compares the average predicted probability for cases and non-cases in the old and new models [66]. The NRI and IDI were calculated to assess model improvement following the inclusion of genetic liability in the models. This was implemented using the *“reclassification”* function from the *PredictABEL* packages in the R-program. Higher IDI value indicated better discrimination and higher NRI value indicated better risk reclassification by the new model [67–69]. The above performance metrics have been discussed in detail elsewhere [64] and in our previous work [33].

## 5.0 Results

### 5.1 Study Characteristics

Table 1 presents the baseline characteristics of the study. The study included 243,339 unrelated UK Biobank participants of European ancestry. The average age of participants included in the study was 55.4 (SD=7.98) years at recruitment. Over half of the sample were women (N = 141,212; 58%). During a median follow-up of 8.22 years, 1,973 first ever stroke episodes of which 45.3% were women were recorded among the participants.

**Table 1:** Baseline Characteristics of Study Population stratified for stroke event and non-stroke event Within the UK Biobank Population.

| **Characteristic** | **Overall**<br>**(N=243,399)** | **Non-event**<br>**(N=241,426)** | **Stroke event**<br>**(N=1973)** | **HR (95% CI)** | **P-value** |
| --- | --- | --- | --- | --- | --- |
| **DM, Yes; n (%)** | 6939 (2.9%) | 6826 (2.8%) | 113 (5.7%) | 2.08(1.72, 2.51) | <0.001 |
| **Hypertension, Yes;**<br>**n (%)** | 116216 (47.7%) | 114840 (47.6%) | 1376 (69.7%) | 2.52(1.29, 2.78) | <0.001 |
| **Sex, Male; n (%)** | 102187 (42.0%) | 101107 (41.9%) | 1080 (54.7%) | 1.67 (1.53, 1.83) | <0.001 |
| **Age (Years), Mean**<br>**(SD)** | 55.4 (7.98) | 55.4 (7.98) | 60.0 (7.14) | 1.93 (1.83, 2.03) | <0.0001 |
| **Body Mass Index**<br>**(kg/m<sup>2</sup>), Mean**<br>**(SD)** | 26.8 (4.57) | 26.8 (4.57) | 27.4 (4.83) | 1.12 (1.08, 1.17) | <0.001 |
| **Total Cholesterol**<br>**(mmol/l), Mean**<br>**(SD)** | 5.91 (1.06) | 5.91 (1.06) | 5.94 (1.09) | 1.03 (0.98, 1.07) | 0.30* |
| **LDL (mmol/l),**<br>**Mean (SD)** | 4.68 (2.37) | 4.67 (2.36) | 5.03 (2.51) | 1.03 (1.03, 1.12) | 0.002 |
| **Smoking** |  |  |  |  |  |
| Current; n (%) | 76397 (31.4%) | 75647 (31.3%) | 750 (38.0%) | REF | REF |
| Previous; n(%) | 2900 (1.2%) | 2855 (1.2%) | 45 (2.3%) | 1.58 (1.17, 2.13) | 0.003 |
| Never; n (%) | 164102 (67.4%) | 162924 (67.5%) | 1178 (59.7%) | 0.73 (0.67, 0.80) | <0.001 |
| **Alcohol** |  |  |  |  |  |
| Current; n(%) | 228349 (93.8%) | 226556(93.8%) | 1793 (90.9%) | REF | REF |
| Previous; n(%) | 7082 (2.9%) | 6996 (2.9%) | 86 (4.4%) | 1.55 (1.25, 1.93) | <0.001 |
| Never; n (%) | 7968 (3.3%) | 7874 (3.3%) | 94 (4.8%) | 1.50 (1.22, 1.85) | <0.001 |

\*The P-value is from a univariate analysis of the Cox Proportional Hazard Model comparing the distribution of the baseline characteristics among stroke event and non-stroke event. \*Not significant; DM= Diabetes Mellitus, HR = Hazard Ratio, CI = Confidence Interval, REF: Reference.

In the overall sample, 76,397 participants (31.4%) were current smokers, and 228,349 participants (93.8%) were current alcohol drinkers. All the conventional risk factors included in the analysis showed a statistically significant association with the risk of incident stroke in univariate analysis except total cholesterol (Table **1**). The prevalence of DM within the sample was 2.9% (N = 6,939) while hypertension was 47.8% among the participants (N = 116,216). The univariable Cox association analysis results indicated that age, sex, BMI, hypertension, DM, LDL, alcohol use, and smoking history were statistically associated with risk of stroke (**Table 1**). These variables were used as the features to construct conventional risk factors models. The correlation matrix (**Figure 2**) between the characteristics in the study demonstrated that total cholesterol and LDL were highly correlated (r^2^ = 0.94). LDL was used in further analysis and feature selection.

**Figure 2.**
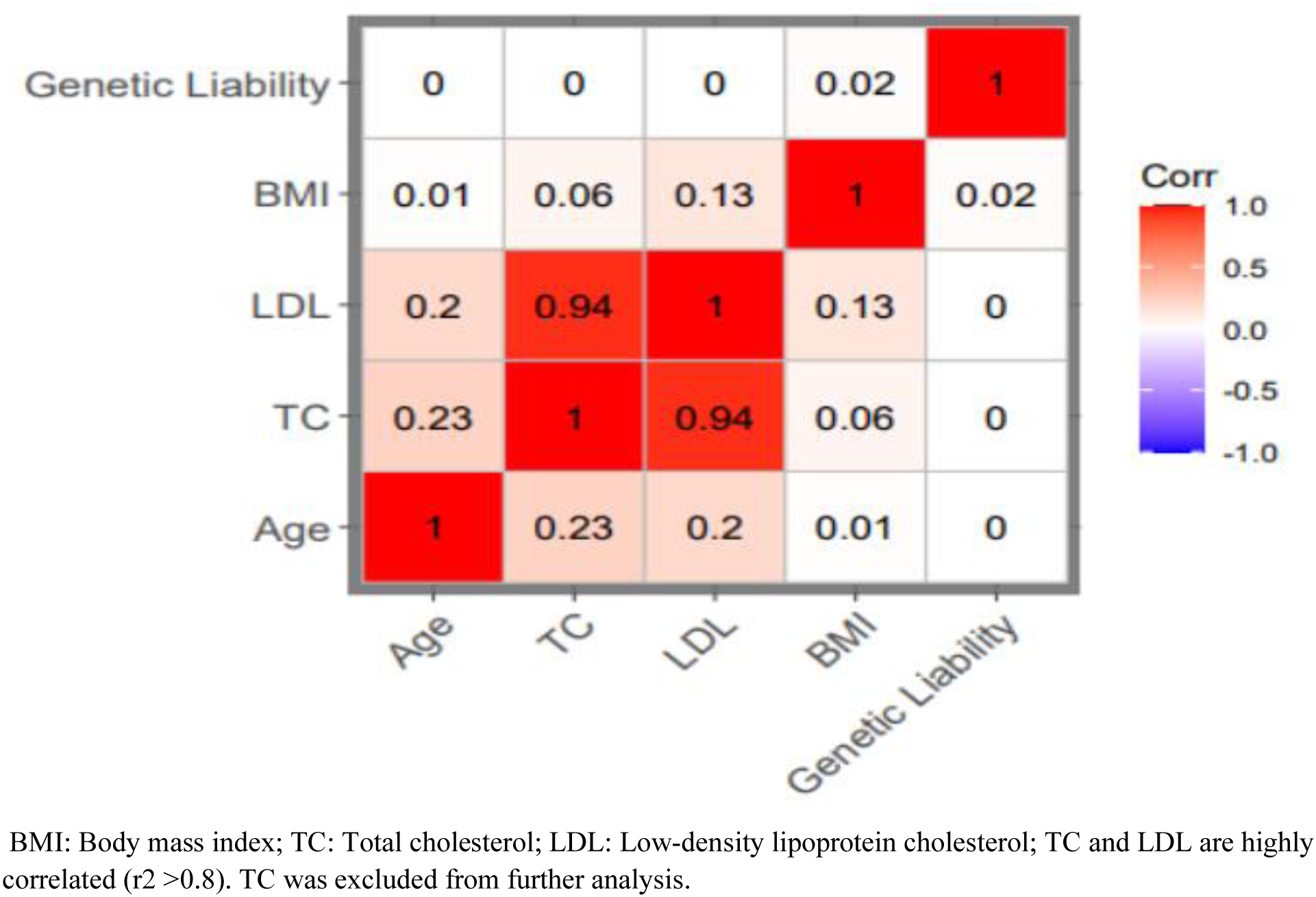
Correlation matrix plot: The plot shows the correlation coefficients between numerical features. TC and LDL are highly correlated (r^2^ >0.8). LDL was excluded from further analysis (prediction model construction). BMI: Body mass index; TC: Total cholesterol; LDL: Low-density lipoprotein cholesterol.

### 5.2 The Relationship Between Genetic Liability and Stroke

Kaplan–Meier curve showed differences in stroke incidents and cumulative hazard between the high risk and low risk genetic liability groups (**Figure 3).** Each unit (standard deviation) increase in genetic liability increases the risk of incident stroke by 7% (HR = 1.07, 95% CI = 1.02, 1.12, P-value = 0.003; **Table 2**; **Figure 4**).

**Figure 3:**
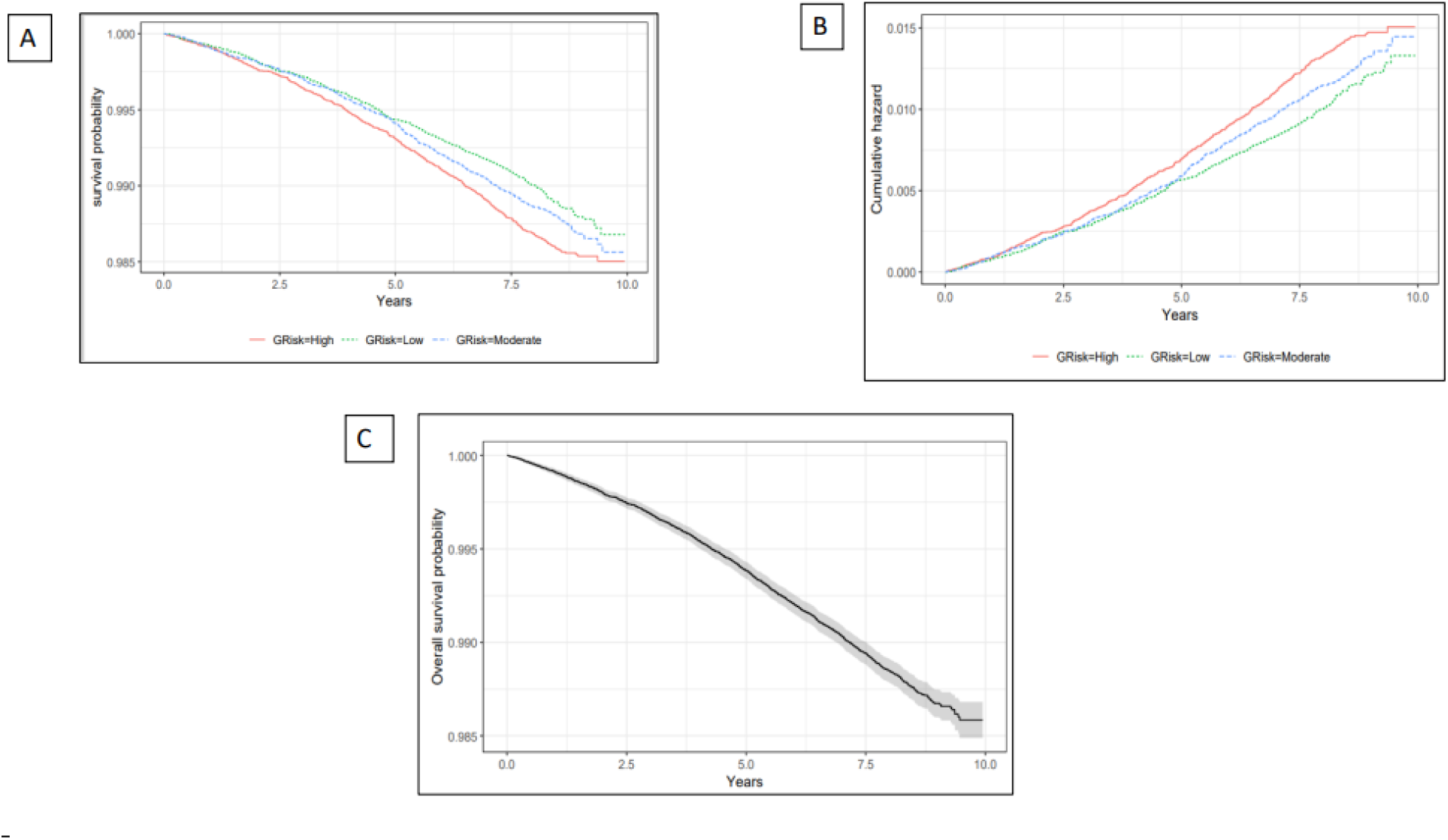
Survival probability and Cumulative hazard plot stratified by genetic risk level: Panel A. Survival probability plot stratified by genetic risk level. Panel B. Cumulative hazard plot stratified by genetic risk level. Panel C. Overall survival probability of the study population. Panels A and B demonstrate the difference in risk of stroke between genetic liability categories. Panel C: As years go by, the risk of stroke increases.

**Figure 4:**
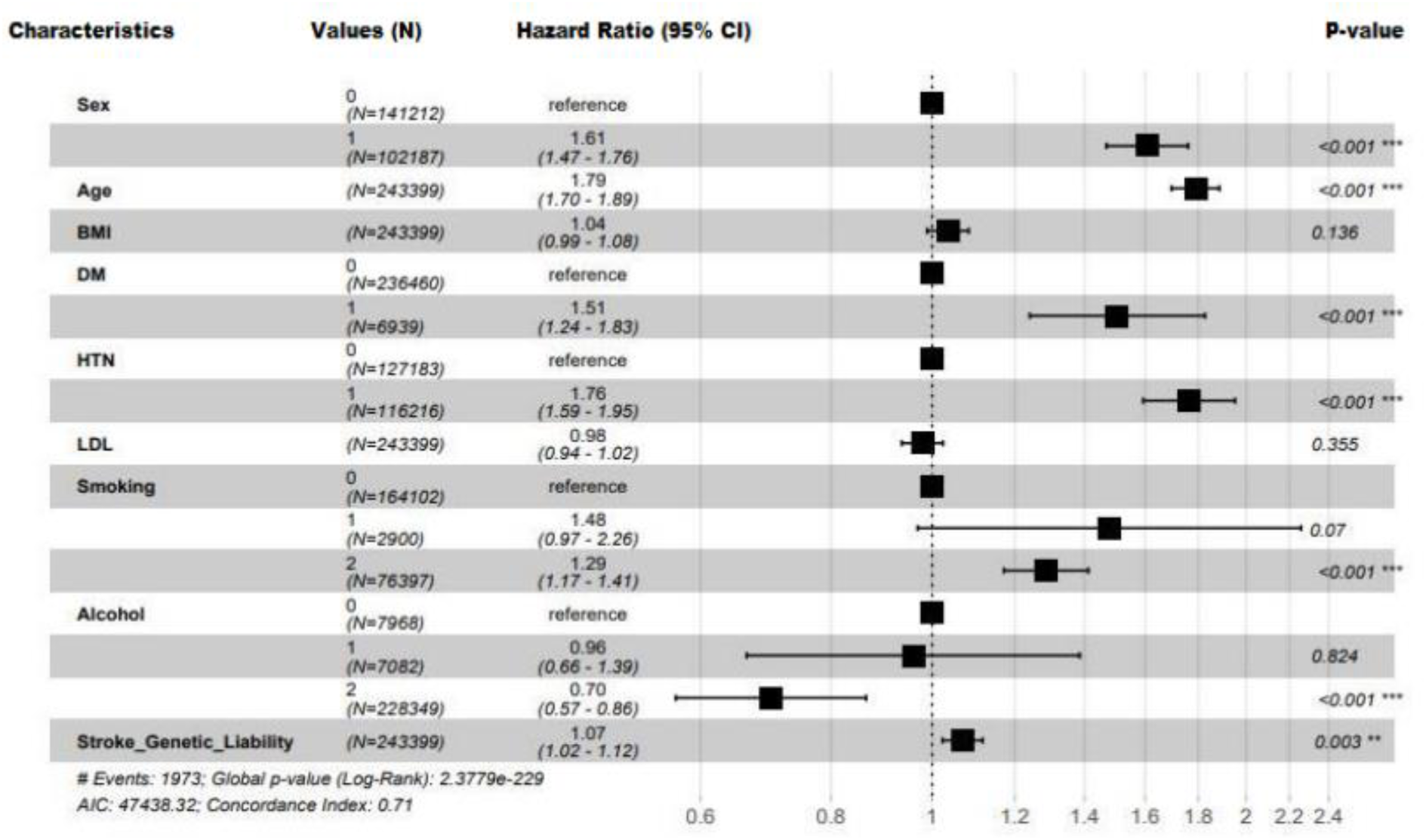
Forest plot of the full Cox proportional hazard model: The vertical line at Hazard ratio (HR) = 1 acts as the null hypothesis reference. The horizontal line represents the confidence interval (CI). If the CI of a predictor does not cross the vertical line, then the HR of that predictor is statistically significant (P-value < 0.05) otherwise HR is statistically insignificant (P-value > 0.05).

**Table 2:** The Result of Univariable Cox Proportional Hazard Model for The Association of Genetic Liability (Categorical and Continuous) With Incident Stroke Within the UK Biobank Population.

| **Genetic liability Level** | **HR (95% CI)** | **P-value** | **HR (95% CI)** | **P-value** | **HR (95% CI)** | **P-value** | **HR (95% CI)** | **P-value** |
| --- | --- | --- | --- | --- | --- | --- | --- | --- |
|  | **Model 1** |  | **Model 2** |  | **Model 3** |  | **Model 4** |  |
| Moderate Risk | 1.06(0.95, 1.18) | 0.31 | 1.06(0.95, 1.18) | 0.31 | 1.05 (0.94, 1.17) | 0.04 | 1.05(0.94, 1.17) | 0.40 |
| High Risk | 1.15(1.03, 1.28) | 0.01 | 1.16(1.04, 1.30) | 0.01 | 1.14(1.02, 1.27) | 0.02 | 1.14(1.02, 1.27) | 0.02 |
| **Genetic liability (continuous)** | 1.08(1.03, 1.13) | <0.001 | 1.08(1.03,1.13) | <0.001 | 1.07(1.03, 1.12) | 0.002 | 1.07(1.02, 1.12) | 0.003 |

Model 1: univariable Cox proportional hazard. Model 2: Adjusted for age and sex. The low genetic risk group was considered as the reference. Model 3: Adjusted for Age, Sex, BMI, LDL, HTN, and DM. Model 4: Adjusted for Age, Sex, BMI, LDL, HTN, DM, Smoking status, and Alcohol status. BMI: Body mass index; LDL: Low-density lipoprotein cholesterol; HTN: Hypertension; DM: Diabetes mellitus.

The risk of stroke was greater in the higher genetic liability group demonstrated by a 14 % increased risk (HR = 1.14, 95% CI = 1.02, 1.27, P-value = 0.02) compared with the low genetic liability group. The global Schoenfeld P-value from the Schoenfeld test (P-value = 0.14; **Table 4**) indicating the proportional hazard (PH) assumption is reasonable for the model (**Supplementary Figure 3**).

### 5.3 Prediction Value of the Conventional Factors

**Table 3** summarizes the performance of the prediction models in the testing set. We considered prediction only up to the median follow up time of 8.22 years.

**Table 3:** Assessment of the proportional hazard (PH) assumption using the global Schoenfield test.

| Characteristics | chisq | df | P-value |
| --- | --- | --- | --- |
| Sex | 0.42 | 1 | 0.52 |
| Age | 0.82 | 1 | 0.37 |
| BMI | 7.49 | 1 | 0.01 |
| DM | 0.08 | 1 | 0.78 |
| HTN | 0.14 | 1 | 0.71 |
| LDL | 0.36 | 1 | 0.55 |
| Smoking | 1.40 | 1 | 0.24 |
| Alcohol | 0.12 | 1 | 0.73 |
| SGL | 2.48 | 1 | 0.12 |
| GLOBAL | 13.43 | 9 | 0.14 |

This table illustrates an assessment of the proportional hazard (PH) assumption using the global Schoenfeld test that assesses proportional hazard assumption for all covariates from a multivariate model. The test indicated a p-value of 0.14 indicating no significant time-dependent effect on the covariates jointly. SGL: Stroke genetic liability; BMI: Body mass index; LDL: Low-density lipoprotein cholesterol, DM: Diabetes mellitus; HTN: Hypertension; df::Degree of freedom, se: standard error; chisq: chi-square.

**Table 4:** The Result of prediction value of stroke genetic liability score in for incident stroke in the UKB.

|  | Models | AUC 95%CI | NRI (95% CI) | P-value for NRI | IDI (95% CI) | P-value for IDI | Brier Score | ICI |
| --- | --- | --- | --- | --- | --- | --- | --- | --- |
| **Coxph** | Model 1 | 69.43<br>(67.30, 71.56) | REF | REF | REF | REF | 0.01 | 0.002 |
| | Model 2 | 69.54<br>(67.40, 71.68) | 0.20<br>(0.119, 0.285) | 0.00 | $1.0 \times 10^{-04}$<br>(0.000, $3.0 \times 10^{-04}$) | 0.14 | 0.01 | 0.002 |
| **GBM** | Model 1 | 69.34<br>(67.23, 71.50) | REF | REF | REF | REF | 0.01 | 0.001 |
| | Model 2 | 69.38<br>(67.26, 71.50) | -0.11<br>(-0.193, -0.027) | 0.01 | 0.00<br>($-1.0 \times 10^{-04}$, $1.0 \times 10^{-04}$) | 0.61 | 0.01 | 0.001 |
| **DT** | Model 1 | 67.58<br>(65.46, 69.70) | REF | REF | REF | REF | 0.01 | 0.001 |
|  | Model 2 | 67.58<br>(65.46, 69.70) | 0.00<br>(0.00, 0.00) | NaN | 0.00<br>(0.000, 0.000) | NaN | 0.01 | 0.001 |
| **RF** | Model 1 | 65.62<br>(63.48, 67.75) | REF | REF | REF | REF | 0.01 | 0.003 |
| | Model 2 | 65.35<br>(63.18, 67.52) | 0.17<br>(0.087, 0.249) | $5.0 \times 10^{-05}$ | 0.00<br>($-7.0 \times 10^{-04}$, $8.0 \times 10^{-04}$) | 0.98 | 0.01 | 0.003 |

Model 1 (Basic model) features: Age, Sex, BMI, HTN, DM, LDL, Smoking status, and Alcohol status. Model 2 features: Age, Sex, BMI, HTN, DM, LDL Smoking status Alcohol status and genetic liability. BMI: Body mass index; LDL: Low-density lipoprotein cholesterol; HTN: Hypertension; DM: Diabetes mellitus. ICI: Integrated calibrated index; REF: Reference; NRI: Continuous Net Reclassification Index; IDI: Integrated Discrimination Index; NaN: Not a Number. Calibration metric (ICI) is based on a calibration curve estimated for a time-to-event outcome (time = 8.20 years) via a restricted cubic spline using complementary log-log transformed predicted probabilities with the “*pmcalibration*” function in the R program.

The Cox Proportional Hazard model with the conventional risk factors (the model without genetic liability) showed a moderate performance and discrimination (AUC= 69.43; 95% CI = 67.30, 71.56; BS = 0.01, and ICI = 0.002) compared with the Gradient Boosting Machines approach (AUC=69.34 ; 95% Cl = 67.23, 71.50; BS = 0.01, and ICI = 0.001), the Decision Tree models (AUC= 67.58;95% CI = 65.46, 69.70, BS = 0.01, and ICI = 0.001), and the Random Forest model which showed the lowest performance (AUC=65.62;95% CI = 65.48, 67.55, BS = 0.01, and ICI = 0.003).

### 5.4 Prediction Value of Genetic Liability

The prediction value of the Cox proportional hazards model improved slightly when the stroke genetic liability was incorporated into the model with conventional risk factors (AUC = 69.54; 95% CI = 67.40, 71.68; AUC change=0.16%; **Table 3**). We also observed a slight improvement in risk reclassification, leading to an overall NRI value of 0.20 (95% CI = 0.119, 0.285; P-value = 0.00; **Table 3**). The IDI value of Cox proportional hazard was negligibly improved by 1.0×10^-04^ (95% CI = 0.000, 3.0×10^-04^; P-value = 0.14; **Table 3**).

The Gradient Boosting Machine model slightly improved in prediction performance (AUC = 69.38; 95% CI = 67.26, 71.50; **Table 3)** but deteriorated in NRI by a value of -0.11 (95% CI = - 0.193, -0.027; P-value = 0.01; **Table 3**) after adding the stroke genetic liability. There was no improvement in overall IDI value using any of the machine learning models.

Using Decision Tree (AUC= 67.58, 95% Cl = 65.46, 69.70, BS = 0.01, and ICI = 0.001) or Random Forest (AUC= 65.35; 95% CI = 65.48, 67.55, BS = 0.01, and ICI = 0.003), no improvement in prediction performance was observed adding genetic liability (**Table 3)**. The overall NRI for Random Forest was improved by NRI = 0.17 (95% CI = 0.087, 0.249; P-value = 5.0×10^-05^; **Table 3**) but not for the Decision Tree technique.

## 6. Discussion

### 6.1. Main Findings

The present study included genome-wide stroke genetic liability (using 252,903 genetic variants) for 243,399 participants of European descent over a median follow-up of 8.22 years. Our findings indicate that (1) the genome-wide stroke genetic liability is independently associated with the risk of stroke, (2) A prediction model integrating the genome-wide stroke genetic liability provides a slight improvement in prediction performance beyond the conventional risk factor for stroke, and (3) the Cox proportional hazard method showed better prediction performance than machine learning models (Random Forest, Gradient Boosting Machines, and Decision Tree) with or without incorporation of genetic liability in the model.

Our first finding that stroke genome-wide genetic liability increases risk of stroke is consistent with previous studies [22–26, 29] including studies by Myserlis *et al*. [22], Rutten-Jacobs *et al. al*. [23], Yang *et al.* [24], Abraha*m et al*. [25], Verbaas *et al*. [26] and Hachiya [29] that reported that stroke genetic liability is a strong independent predictor of risk of future stroke occurrences.

These previous studies mainly calculated stroke genetic liability based on a limited selection of single-nucleotide polymorphisms (SNPs) that have strong associations with the traits. Our result is a step forward in the sense that we present the risk of stroke imposed by a whole genome genetic liability of stroke in a European setting. Yang *et al.* [24] estimated a whole genome genetic liability of stroke (stroke and its subtypes) in China Kadoorie Biobank and showed that genetic liability of stroke increases risks of any stroke (14%), ischemic stroke (7%), and intracerebral hemorrhage (10%). We observed a 15% greater risk of any stroke among European participants with a high genome-wide stroke genetic liability compared with those with a low genetic liability which is comparable to the study by Yang *et al.* [24] in a Chinese population. Our study also differs from previous studies, including the definition or classification of the outcome and sample characteristics. We defined stroke events as any cases of (1) ischemic stroke, (2) intracerebral hemorrhage, (3) subarachnoid hemorrhage, (4) other cerebrovascular disease, or (5) stroke that is not specified as hemorrhage or infarction. We thus captured a broader definition of stroke, which could have increased the stroke diversity in our analysis. To investigate the relationship between genetic liability and stroke, Rutten-Jacobs et al. [23] generated a genetic liability from 90 SNPs associated with stroke (at a P-value less than 1×10^−5^). They demonstrated a 7 to 13% increase in the risk of stroke for each standard deviation increase in the genetic liability. Myserlis *et al.* [22] and Abraham *et al.* [25] included genetic liability of stroke within a meta-scoring technique that combined 19-21 distinct genetic liabilities to form a metaGRS. These studies found that the metaGRS was associated with an increased risk of incidence of intracerebral hemorrhage [22] and ischemic stroke [25]. They showed a 15% increase in the risk of intracerebral hemorrhage and a 26% increase in the risk of ischemic stroke for each standard deviation increase in the metaGRS. The association was stronger than any of the individual genetic liabilities included in the metaGRS. However, their results did not distinguish the effect of genetic liability of stroke per se as it was part of a MetaGRS comprising 19-21 distinct genetic liabilities for various traits. Abraham *et al.* included several genetic liabilities for multiple stroke-related phenotypes including ischemic stroke, any stroke, small vessel stroke, large artery stroke, cardioembolic stroke, and several stroke risk factors in their metaGRS. Myserlis *et al.* included genetic liabilities for multiple phonotypes including white matter hemorrhage (n=87,951 SNPs) and small vessel stroke (n=2,162 SNPs) within the metaGRS calculation. While metaGRS has been found to improve risk prediction, there may be some biases in prediction performance because it was built using elastic-net regression. Additionally, certain SNPs included in the calculation of individual phenotypes’ genetic liabilities may be associated with several phenotypes [26]. Therefore, the metaGRS may contain overlapping information due to possible correlation among the genetic liabilities included in the metaGRS [70]. Our approach to considering genome-wide genetic liability for stroke aimed to capture the polygenic component of stroke i.e., we had no threshold for selection of SNPs associated with stroke. Thus, we included all SNPs even those with small or non-significant effects. It is known that this approach would increase the accuracy of effect estimated for genetic liability and therefore improve accuracy in the identification of high-risk individuals [71, 72].

Our prediction models demonstrated that the genome-wide stroke genetic liability may slightly enhance (1) overall stroke prediction performance to distinguish the cases (the Cox proportional hazards model and the Gradient Boosting Machine) and (2) correct classification of individuals at risk beyond conventional risk factors (the Cox proportional hazards model and Random Forest). However, none of our models demonstrated statistically improved predicted probabilities for cases and non-cases based on IDI.

Our findings are supported by the results reported in previous studies [40, 73, 74] observing that adding genetic liability and conventional risk factors in risk prediction models improves discrimination performance compared to using only conventional risk factors. Papadopoulou *et al.* [40] used genetic liability based on 28 SNPs in a European population focused on ischemic stroke in patients with atrial fibrillation (AF) and observed that XGBoost performed better than the CHA2DS2-VASc model, an existing clinical model for calculating stroke risk for patients with atrial fibrillation. Cárcel-Márquez *et al*. [73] used genetic liability based on 93 SNPs to predict cardioembolic stroke in the European population using logistic regression while Jung et al. [74] used genetic liability based on 16 SNPS in an Asian population to predict stroke in the Korean population using Cox proportional hazards regression. The prediction performance of our best model was higher than that reported by both Papadopoulou *et al.* [40] and Jung *et al*. [74] but lower than the results reported by Cárcel-Márquez *et al.* [73]. It should be noted that the results reported by Cárcel-Márquez *et al.* did not use time-to-event data and were focused on prevalent cases of stroke. Whereas our study reports the results for incident stroke.

Our study indicated that the Cox proportional hazard regression models outperformed all the machine learning models in the context of time-to-event data for stroke. Unlike our study, previous studies suggest that machine learning algorithms outperform traditional statistical approaches in the prediction of stroke [32, 41, 42]. This could be due to a small number of events (1,973 stroke events) and a few predictors (up to 9 predictors). Both factors are considered as reasons for Cox models to outperform machine learning [75] implying that no expectations should be made for machine learning models to always outperform Cox models.

We found that genetic liability improved stroke risk classification for less than 1% of the subjects. Health economy studies could consider investigating if using this information in the identification of high-risk individuals to target for stroke prevention programs could make a significant cost-effective change in stroke related expenses.

The large sample size of UK Biobank and the number of incident strokes enabled the statistical power for our analysis in which we used time-to-event data for over 200,000 individuals of European ancestry, with a median follow-up of 8.22 years. A distinctive feature and the strength of our study compared with previous studies is that we generated genetic liability for stroke using over 250,000 genetic variants.

Validation in external cohort datasets could improve the precision of our findings. To minimize lack of validation in external cohorts, we internally validated our machine learning models in the testing set, where we randomly partitioned data into a training set (70% of participants) for developing the prediction models and a testing set (30% of participants) to evaluate the prediction models’ performance.

### Conclusions

In conclusion, incorporating genetic liability into stroke risk prediction models could slightly improve prediction performance and should be considered when predicting the risk of stroke. Cox proportional hazard models should be given priority over machine learning models in the prediction of the risk of stroke.

## Data Availability

All data produced in the present study are available upon reasonable request to the authors

## Acknowledgment

This research has been conducted using the UK Biobank Resource under Application Number 60549. The MEGASTROKE project received funding from sources specified at http://www.megastroke.org/acknowledgments.html".

## Author contributions

Conceptualization, R.P.; Data curation, Formal analysis, G.M.; Investigation, G.M.; Methodology, G.M., R.P.; Project administration, G.M., and R.P.; Resources, R.P.; Supervision, R.P; Writing— original draft, G.M.; Writing—review & editing, G.M., and R.P. All authors have read and agreed to the published version of the manuscript.

## Institutional Review Board Statement

The study was conducted in accordance with the Declaration of Helsinki and approved by the Institutional Review Board (or Ethics Committee) of Brunel University London, College of Health, Medicine, and Life Sciences (27684-LR-Jan/2021-29901-1).

## Informed Consent Statement

Informed consent was obtained from all subjects involved in the study.

## Data Availability Statement

Not applicable.

## Supplementary Materials

**Supplementary Data S1-** Supplementary Data S1: List of genetic variants summary statistics used to construct the genetic risk scores.

