## Supplementary Figures for "Evaluation of Machine Learning and Traditional Statistical Models to Assess the Value of Stroke Genetic Liability for Prediction of Risk of Stroke within the UK Biobank"

**Supplementary Figure 1: Overview of the process to create genetic liability for stroke within the UK Biobank**

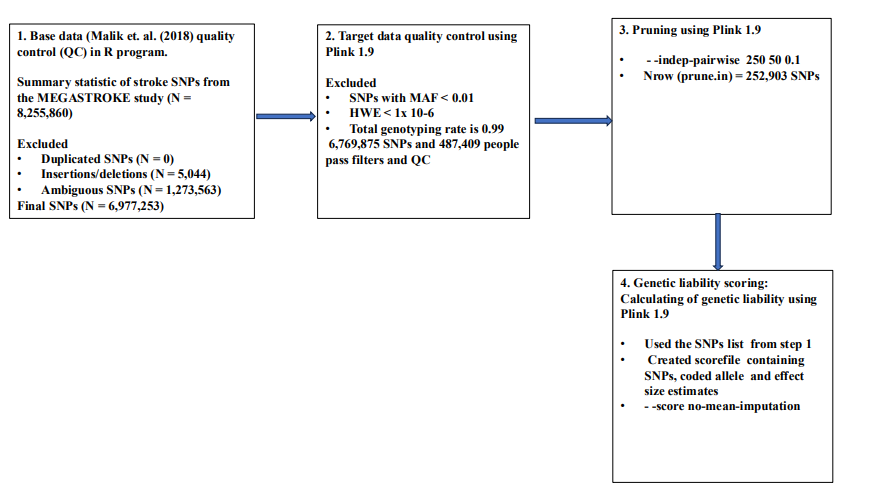
**Genetic liability calculation process: SNP**: Single nucleotide polymorphism; **MAF**: Minor allele frequency; **LD**: Linkage disequilibrium; **HWE**: Hardy-Weinberg Equilibrium. The SNPs were pruned with plink command - -indep-pairwise window size = 250, step size = 50, r^2^ = 0.1. **Base data**: SNP list from Malik et. al. (2018), **Target data**: genotype data in plink binary format.

**Supplementary Figure 2: Overview of the modeling and predictions process for using machine learning and genome-wide genetic liability for prediction of risk of stroke within the UK Biobank.**

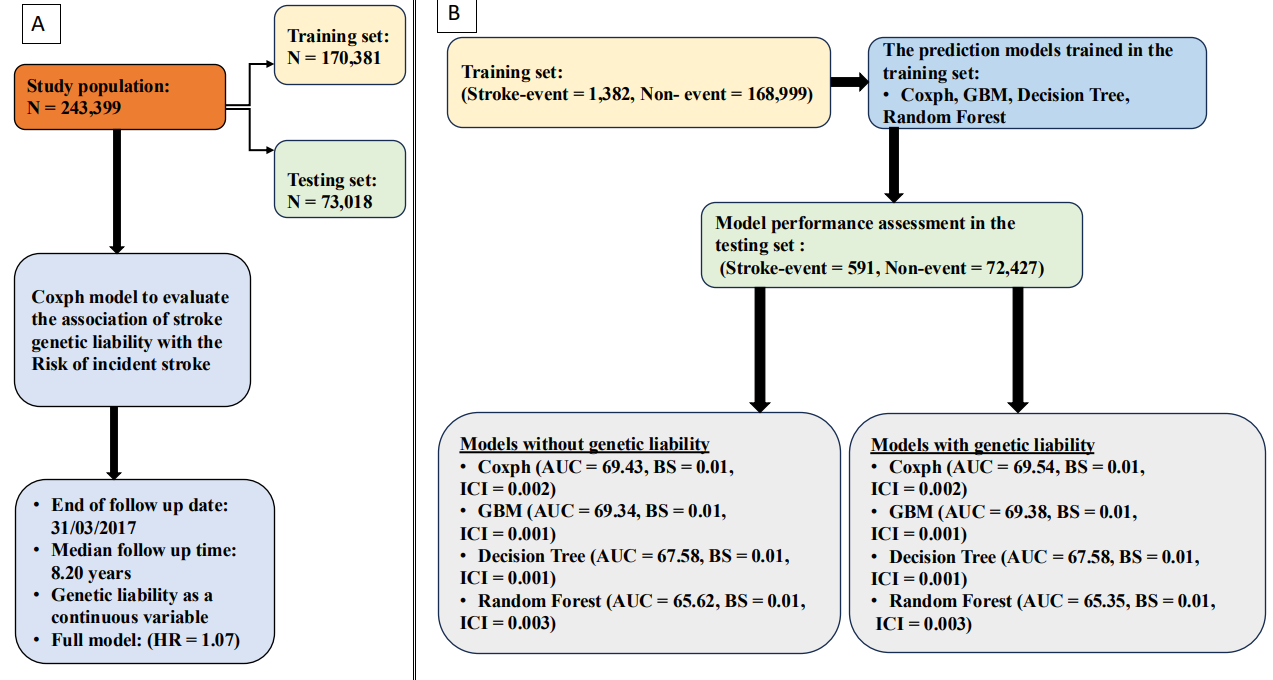
**Stroke risk prediction model creation and performance evaluation:** Coxph: Cox proportional hazard, GBM: Gradient Boosting model, HR= Hazard ratio, AUC: Area under the curve, BS: Brier score, ICI: Integrated calibrated index. Panel A: Assessing the association between genetic liability and incident stroke. Panel B: Stroke risk prediction modeling and performance evaluation.

**Supplementary Figure 3: Schoenfield test results of full Cox proportional hazard model.**

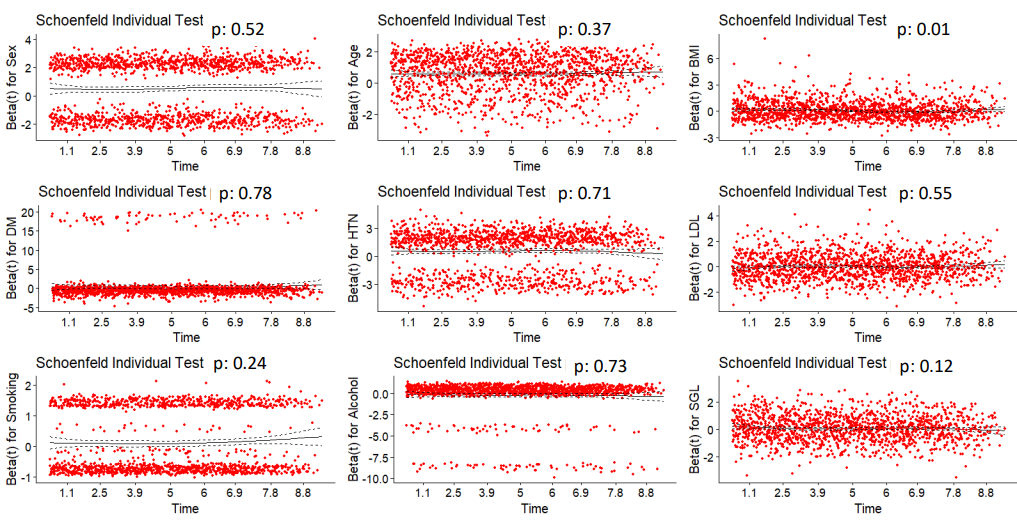
The figure illustrates an assessment of the proportional hazard (PH) assumption using the global Schoenfield test that assesses proportional hazard assumption for all covariates from a multivariate model. The test indicated a p-value of 0.14 indicating no significant time-dependent effect on the covariates jointly. If P-value **>** 0.05, the test fails to reject the null hypothesis i.e. the PH assumption would hold for the overall model and covariates have a consistent effect over time. The individual Schoenfield test for BMI indicates that variable does not have a consistent effect over time. Thus, BMI is adjusted within all the Cox models in the study. BMI: Body Mass Index, LDL: Low-density lipoprotein cholesterol, SGL: Stroke genetic liability

:
